# Reaching young and low-income people for sexual healthcare – perspectives from healthcare workers: a qualitative study

**DOI:** 10.1101/2025.09.25.25332954

**Authors:** Leonie Arns-Glaser, Elea Widrig, Vasileios Nittas, Jan S. Fehr, Benjamin Hampel, Andrea Farnham

## Abstract

**Background:** Free voluntary counselling and testing (VCT) interventions improve access to HIV and sexually transmitted infections (STI) prevention, particularly for underserved populations. In 2023, Zurich (Switzerland) introduced a pilot programme offering free VCT for young people and low-income residents. This study explores healthcare professionals’ (HCPs) experiences implementing the programme, focusing on perceived benefits, barriers, and opportunities for improvement.

**Methods:** We conducted a qualitative descriptive study with 12 HCPs from two participating clinics. Data collection included semi-structured interviews and semi-quantitative ranking activities to prioritise perceived barriers and hardly reached population groups were analysed using inductive thematic analysis and data frame comparison. Standards for reporting qualitative research guidelines (SRQR) informed reporting.

**Results:** HCPs viewed the programme as an effective entry point into sexual healthcare for young people and first-time testers, particularly due to the removal of financial barriers. Counselling was seen as its most important component, promoting knowledge-sharing and early engagement in preventative healthcare. The programme successfully reached well-informed, highly educated individuals. However, key high-risk groups - including vocational students, conservative individuals, asylum seekers, and those under 18 – were not well reached. Reported barriers included stigma, misinformation, and long appointment waiting times. HCPs recommended improved outreach to vocational schools, more inclusive educational materials, and mobile testing initiatives to improve accessibility.

**Conclusion:** HCPs consider free VCT programmes a critical tool for equitable access to sexual healthcare. However, to maximise their reach and impact, targeted, context-sensitive strategies are needed to engage underserved and high-risk populations more effectively.

**WHAT IS ALREADY KNOWN ON THIS TOPIC:** Financial barriers reduce uptake of HIV and STI testing, particularly among young and low-income populations in high-income countries.

**WHAT THIS STUDY ADDS:** Removing cost improves uptake and visibility of voluntary counselling and testing (VCT) services, especially among socially connected youth. However, high-need groups such as vocational students, asylum seekers, and individuals from conservative backgrounds remain underreached without targeted outreach.

**HOW THIS STUDY MIGHT AFFECT RESEARCH, PRACTICE OR POLICY:** Findings highlight the need for policies that combine free testing initiatives with inclusive sexual health education and outreach to improve equity and programme effectiveness.

## Background

Voluntary counselling and testing (VCT) is a key strategy in HIV and sexually transmitted infection (STI) prevention. Sexual health counselling has been linked to positive sexual health outcomes among adolescents and emerging adults, including reductions in risky sexual behaviours and increased HIV awareness (1). Early engagement with such services during adolescence shapes long-term health behaviours (2), highlighting VCT’s role in prevention and behavioural changes (1).

Despite these benefits, access to VCT remains uneven in many high-income settings. While fees reduce testing uptake in several European countries (3), even when services are free, concerns about stigma, confidentiality, and fear of disclosure can still limit access, especially in marginalised or rural communities (4,5).

Healthcare professionals’ (HCPs) insights as frontline implementers are critical in identifying and addressing practical and contextual barriers to care. Their understanding of local populations can guide more responsive and culturally sensitive services (5,6). For VCT to be effective, access must be consistently accessible and sustainably integrated into routine care (6). The insights of HCPs are essential for achieving responsive and sustainable sexual healthcare. To our knowledge, no studies have explored HCP perspectives on implementing free HIV/STI testing and counselling in high-income countries.

In the city of Zurich, Switzerland, a pilot project offering free VCT for HIV and STIs was launched to improve access for young people and low-income groups. This initiative represents a targeted effort to reduce financial and structural barriers in a high-income setting, with the potential to inform broader integration of equitable sexual health services (7). This study explores the perspective of HCPs implementing the programme, with a focus on perceived impact, implementation barriers, and opportunities for improving access to sexual healthcare. Specifically, we ask: (1) How do HCPs view the impact and utility of the project for clients? (2) Which populations remain underserved, and why? (3) What strategies could improve outreach to low-income populations at the highest risk of negative sexual health outcomes?

## Methods

### Study design

We conducted a qualitative descriptive study using semi-structured interviews to explore HCPs’ perspectives on the implementation of a free HIV/STI VCT programme in Zurich, Switzerland. This approach was selected to capture the lived experiences of providers and identify practical barriers and facilitators to implementation. Reporting follows the Standards for Reporting Qualitative Research guidelines (SRQR) guidelines (see Supplemental Materials S1) (8).

### Study setting and recruitment

This study was conducted as part of a pilot project providing free VCT for HIV, syphilis, chlamydia, gonorrhoea, and hepatitis C to Zurich residents under 26 years old and low-income individuals (as determined by eligibility for a social discount programme). The service was delivered beginning in June 2023 at two community-based sexual health clinics: Checkpoint Zurich, which is specialised on the LGBTI+ populations, and Test-In, which focuses on the general public. In the first year of the pilot project, the median (IQR) age of the people coming to the free VCT was 24 (22–26) years, 46% were assigned female at birth, the largest demographic group were women having sex with men (43%) (7).

We used a total population sampling approach. All HCPs directly involved in the pilot—either administering VCT or conducting client intake—were invited via email and staff meetings. Participation was voluntary and counted as paid working time.

A total of 12 HCPs participated in the study, with eight based at Checkpoint and four at Test-In. Participants’ median age was 34.5 years (range: 22-52). Nine had a medical background and three a background in social and educational fields. At Checkpoint Zurich, 8 of 16 eligible HCPs (50%) participated, and all 4 HCPs at Test-In (100%) took part, resulting in a sample of 12 participants.

### Data collection

Interviews were conducted one-on-one in private rooms between December 2023 and January 2024 by a single interviewer (LAG), using a semi-structured guide (Supplemental Materials S2). Interviewer’s characteristics and potential influences are listed in the Supplemental Materials S3. On average interviews were 45 minutes (range: 29 – 57 minutes) and were audio-recorded upon participants consent.

Recordings were transcribed verbatim and translated into Standard German using Töggl (töggl.ch), an Automatic Speech Recognition (ASR)-based transcription tool designed for Swiss German dialects.

The interview guide was developed based on literature, project update meetings with stakeholders and HCPs, and piloted with an HCP working in the field. It included open-ended questions on the pilot population, barriers to accessing VCT, perceived usefulness, potential improvements, and implementation challenges. Two semi-quantitative tasks were included, in which participants ranked access barriers and categorised population groups by VCT need; rankings were documented with photographs upon participants’ consent (see Supplemental Material S4).

### Public and Patient involvement and engagement

Patients were not involved in the design, recruitment, or conduct of this study, which focused on HCPs perspectives during the implementation of a policy-driven pilot.

Findings are being shared with stakeholders involved in the programme.

### Data analysis

Data were analysed thematically using an inductive approach. Transcripts were reviewed and corrected in MAXQDA24 (VERBI Software, 2024) (9). Two researchers (EW and LAG) independently coded all transcripts. The final coding framework was developed iteratively, with VN refining themes based on emerging concepts. The revised scheme was then applied by EW and LAG to all transcripts. Data frame analysis was carried out by EW, LAG and AF. Discrepancies in coding and theme interpretation were resolved through discussion. Quotations used in the results section were translated into English and lightly edited to improve readability while preserving the original meaning by LAG (fluent in both languages).

### Ethical considerations

In accordance with the declaration of Helsinki, the study was exempted from formal ethical approval under the Human Research Act review by the Cantonal Ethics Committee of Zurich (Req-2023-00594). All data were securely processed and stored in Switzerland in accordance with data protection regulations. Participants orally consented to participation. Recordings are kept on secured server in Switzerland and will be deleted 10 years after publication. Demographic details of HCPs are saved separately from the interview recordings and transcripts.

## Results

### Perceived benefits and utility of the programme

HCPs viewed the free VCT programme as an effective entry point into sexual healthcare, with many emphasising how cost removal encouraged access among young people who might not otherwise seek testing.

> *“I think it’s a very useful project. I think we reach many, many people, who otherwise would not have come to get tested.”* (Interview 5, Test-In)

Beyond increasing access, HCPs observed that the programme contributed to broader awareness and peer-to-peer information sharing. Word-of-mouth promotion, especially among well-connected groups such as international students and young women, was reported to be a key driver of uptake. Clients often reported discovering the service via Google or social media. In both clinics, HCPs noted that many heterosexual men were encouraged by their female partners to get tested (see Table 1).

**Table 1:**
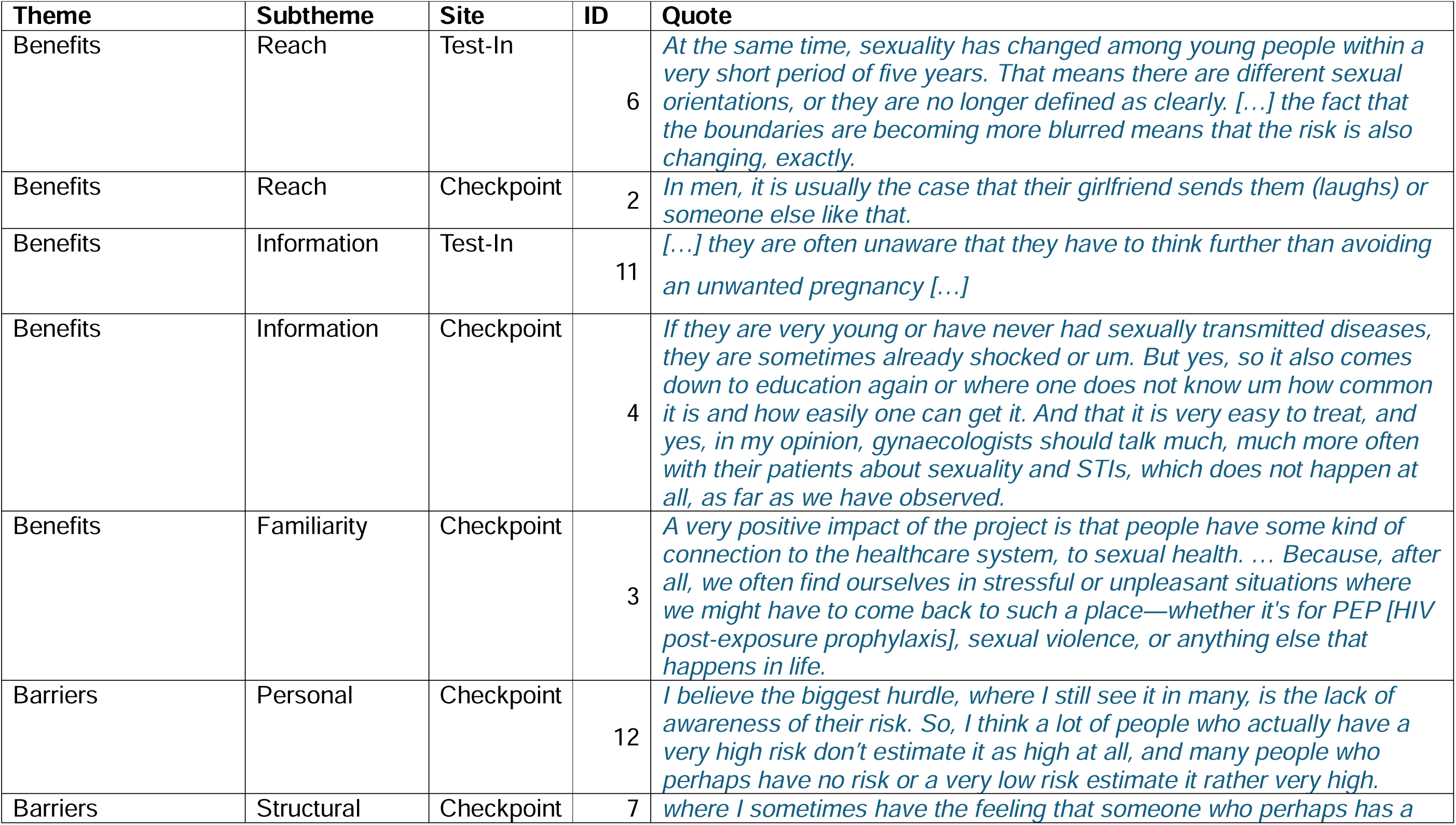
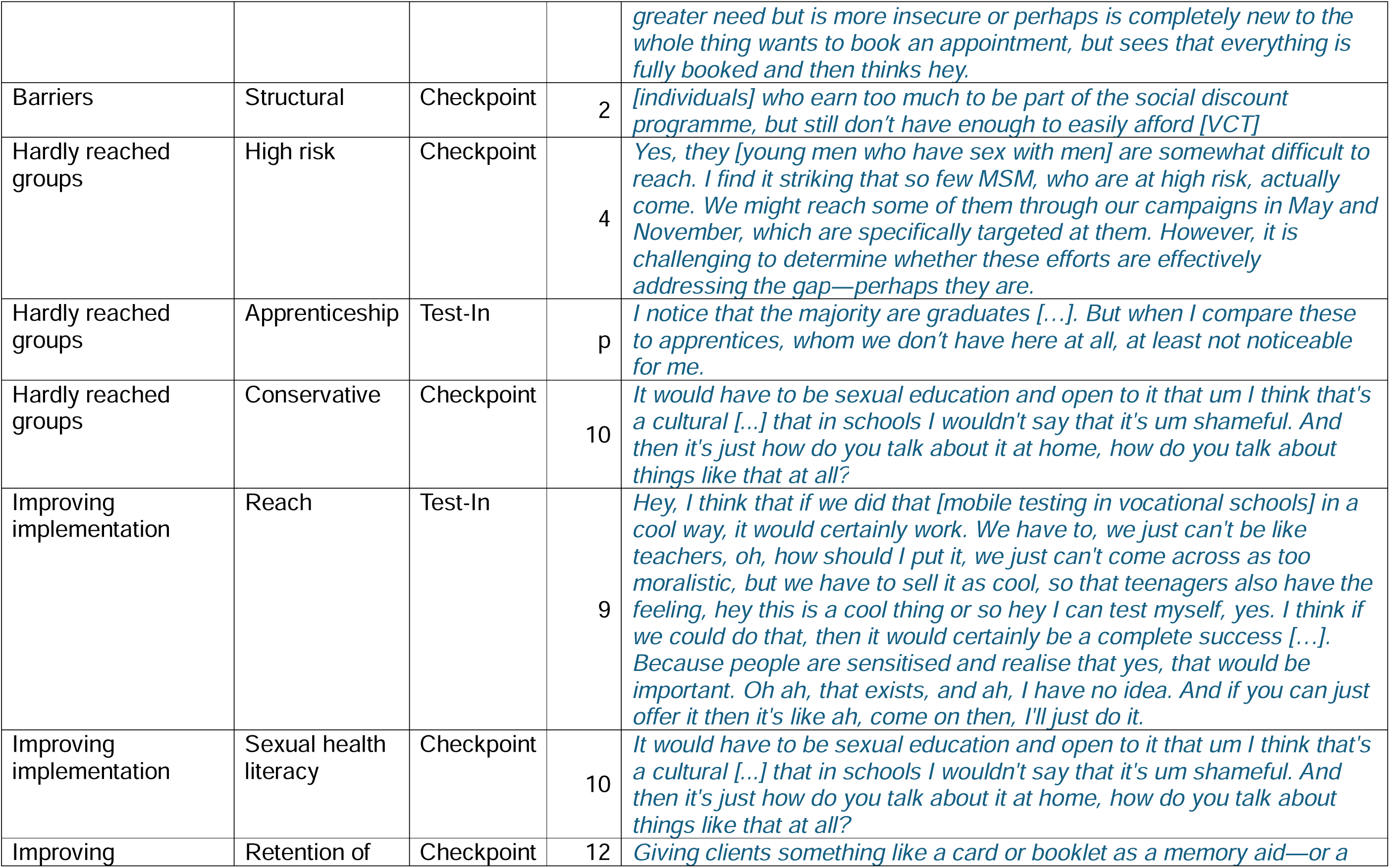
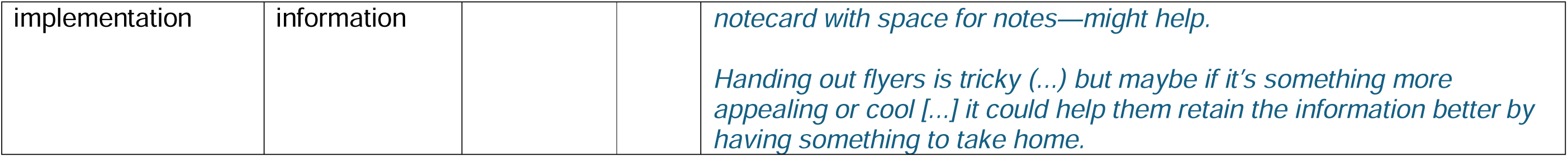
Additional HCP quotes sorted by themes.

| Theme | Subtheme | Site | ID | Quote |
| --- | --- | --- | --- | --- |
| Benefits | Reach | Test-In | 6 | *At the same time, sexuality has changed among young people within a very short period of five years. That means there are different sexual orientations, or they are no longer defined as clearly. [...] the fact that the boundaries are becoming more blurred means that the risk is also changing, exactly.* |
| Benefits | Reach | Checkpoint | 2 | *In men, it is usually the case that their girlfriend sends them (laughs) or someone else like that.* |
| Benefits | Information | Test-In | 11 | *[...] they are often unaware that they have to think further than avoiding an unwanted pregnancy [...]* |
| Benefits | Information | Checkpoint | 4 | *If they are very young or have never had sexually transmitted diseases, they are sometimes already shocked or um. But yes, so it also comes down to education again or where one does not know um how common it is and how easily one can get it. And that it is very easy to treat, and yes, in my opinion, gynaecologists should talk much, much more often with their patients about sexuality and STIs, which does not happen at all, as far as we have observed.* |
| Benefits | Familiarity | Checkpoint | 3 | *A very positive impact of the project is that people have some kind of connection to the healthcare system, to sexual health. ... Because, after all, we often find ourselves in stressful or unpleasant situations where we might have to come back to such a place—whether it's for PEP [HIV post-exposure prophylaxis], sexual violence, or anything else that happens in life.* |
| Barriers | Personal | Checkpoint | 12 | *I believe the biggest hurdle, where I still see it in many, is the lack of awareness of their risk. So, I think a lot of people who actually have a very high risk don't estimate it as high at all, and many people who perhaps have no risk or a very low risk estimate it rather very high.* |
| Barriers | Structural | Checkpoint | 7 | *where I sometimes have the feeling that someone who perhaps has a* *greater need but is more insecure or perhaps is completely new to the whole thing wants to book an appointment, but sees that everything is fully booked and then thinks hey.* |
| Barriers | Structural | Checkpoint | 2 | *[individuals] who earn too much to be part of the social discount programme, but still don't have enough to easily afford [VCT]* |
| Hardly reached groups | High risk | Checkpoint | 4 | *Yes, they [young men who have sex with men] are somewhat difficult to reach. I find it striking that so few MSM, who are at high risk, actually come. We might reach some of them through our campaigns in May and November, which are specifically targeted at them. However, it is challenging to determine whether these efforts are effectively addressing the gap—perhaps they are.* |
| Hardly reached groups | Apprenticeship | Test-In | p | *I notice that the majority are graduates [...]. But when I compare these to apprentices, whom we don't have here at all, at least not noticeable for me.* |
| Hardly reached groups | Conservative | Checkpoint | 10 | *It would have to be sexual education and open to it that um I think that's a cultural [...] that in schools I wouldn't say that it's um shameful. And then it's just how do you talk about it at home, how do you talk about things like that at all?* |
| Improving implementation | Reach | Test-In | 9 | *Hey, I think that if we did that [mobile testing in vocational schools] in a cool way, it would certainly work. We have to, we just can't be like teachers, oh, how should I put it, we just can't come across as too moralistic, but we have to sell it as cool, so that teenagers also have the feeling, hey this is a cool thing or so hey I can test myself, yes. I think if we could do that, then it would certainly be a complete success [...]. Because people are sensitised and realise that yes, that would be important. Oh ah, that exists, and ah, I have no idea. And if you can just offer it then it's like ah, come on then, I'll just do it.* |
| Improving implementation | Sexual health literacy | Checkpoint | 10 | *It would have to be sexual education and open to it that um I think that's a cultural [...] that in schools I wouldn't say that it's um shameful. And then it's just how do you talk about it at home, how do you talk about things like that at all?* |
| Improving | Retention of | Checkpoint | 12 | *Giving clients something like a card or booklet as a memory aid—or a* |
| implementation | information |  |  | *notecard with space for notes—might help.*<br>*Handing out flyers is tricky (...) but maybe if it's something more appealing or cool [...] it could help them retain the information better by having something to take home.* |

HCPs noted gendered patterns in health-seeking behaviour. Women were described as more socialised to attend preventative health visits (e.g., gynaecology visits), whereas heterosexual men lower sexual health literacy and rarely accessed preventative healthcare services. The free VCT programme was seen as a low-threshold opportunity to engage this group.

For many clients, the visit was their first exposure to in-depth, non-judgmental sexual health education. While testing initially drew clients in, the educational discussions enabled knowledge-sharing that extended beyond the individual.

> *“[…] the actual sex education really only happens with us. […] VCT clients are always multipliers as well. […] I always try to give them some sort of ‘go-home message’ that they can hopefully spread […].”* (Interview 12, Checkpoint)

Several HCPs expressed initial scepticism about low-risk clients seeking testing, but most ultimately saw this as a positive opportunity for early engagement and long-term health awareness (see Table 1).

### Populations reached and gaps in reach

HCPs reported that the programme effectively reached students, individuals with university education, and socially connected youth. However, several groups were perceived as underrepresented despite potentially high need.

Young people under 18, particularly those identifying as queer, were rarely seen at Checkpoint, despite the clinic’s traditional focus on LGBTQ+ clients. HCPs speculated that existing targeted programmes for men who have sex with men (MSM) and sex workers may explain their lower-than-expected presence (see Figure 1A).

**Figure 1:**
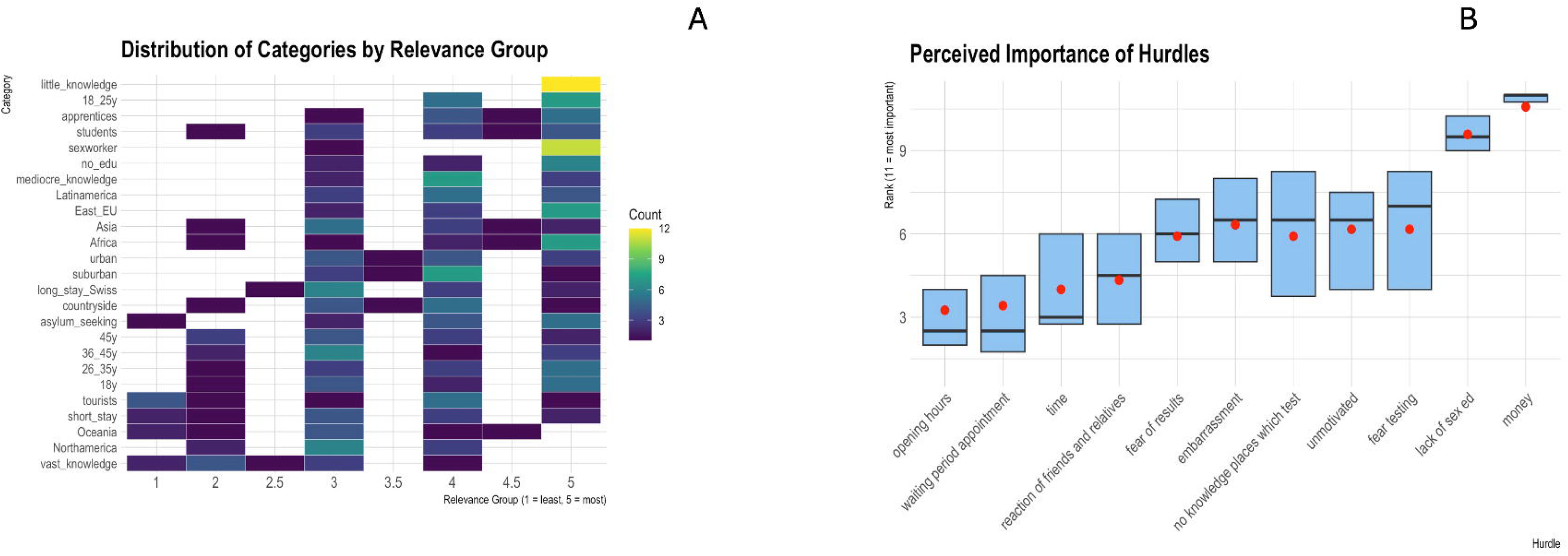
Results of semi-quantitative sorting activities. A shows a heat map in which the sorting of population groups according to their intensity of need for VCT is depicted. 1 = lowest need, 5 = highest need, B depicts the ranking of barriers faced by young people when trying to access VCT prior to the free VCT-programme. The boxplot depicting the median rank with interquartile ranges, whereas the red dot indicates the mean rank.

In contrast, individuals with more conservative worldviews, those not in education or training, and people in low-wage jobs were described as largely absent from the programme. Vocational school students were particularly difficult to reach, with outreach efforts largely unsuccessful.

> *“[…], I wrote to 18 vocational schools […] and I didn’t hear back from any of these 18 vocational schools”* (Interview 9, Test-In)

In the semi-quantitative sorting activity, HCPs ranked asylum seekers, individuals from sub-Saharan Africa, and those with no formal education as the least reached but highest-need groups (see Figure 1A).

### Barriers to access

#### Personal Barriers

HCPs noted that many young people had difficulty accurately accessing their sexual health risks. Some avoided testing due to low perceived risk, while others attended despite very limited sexual experience. This mismatch occasionally led to questions about the programme’s utility but was also viewed as an opportunity for early education.

The semi-quantitative sorting exercise showed that lack of sexual education was the second most important barrier to accessing VCT services after financial concerns. This prioritisation was consistent among HCPs at both sites (see Figure 1B). Other frequently mentioned barriers included embarrassment, fear of a positive result, lack of motivation, and language barriers.

#### Structural Barriers

Structural barriers such as opening hours and long waiting periods were ranked as less critical in the sorting activity, which focused on pre-pilot conditions (see Figure 1B). Nonetheless, several HCPs raised the issue of the current waiting times of up to two months as problematic, particularly for people with urgent needs.

> *“[…] if you have a free offer but no available slots at all, that kind of doesn’t make sense…“* (Interview 7, Checkpoint)

Living with family was another barrier for younger clients, who feared parents discovering testing through insurance or mail correspondence. While universities were receptive to outreach, vocational schools were not. HCPs also mentioned that some individuals not eligible for the social discount programme still faced financial struggles. They observed that individuals qualifying for free testing through the social discount programme were often older students or artists, rather than individuals in low-income employment.

### Recommendations for improving implementation

HCPs suggested a range of strategies to improve reach and impact. These included expanding outreach through mobile testing in locations such schools, bars, clubs, and public spaces where MSM meet for anonymous sex, and creating more anonymous, less queer-coded VCT spaces to attract individuals with more traditional mindsets. Schools were viewed as promising venues for deep prevention among youth, especially those from conservative households (see Table 1).

Additionally, HCPs recommended revising informational materials to be more inclusive of heteronormative populations and women. MSM-focused materials were seen as potentially overwhelming or off-putting to other groups. Several HCPs expressed a desire for shorter appointments or other process improvements to increase daily capacity without compromising quality of care, noting that reducing the initial appointment times by 5 minutes already allowed for one additional appointment per day.

## Discussion

This study explored the perspectives of HCPs involved in the implementation of a free HIV/STI VCT programme in Zurich, Switzerland (7). HCPs generally perceived the programme as highly beneficial in increasing access to sexual health services, particularly in young, well-connected populations. Counselling was viewed as a core component, with potential to improve sexual health literacy and encourage peer knowledge-sharing. However, several high-need populations—including younger adolescents, individuals from conservative or migrant backgrounds, and those in low-income employment—were perceived as underrepresented. Barriers to access included stigma, limited sexual health education, and logistical challenges such as long waiting times.

These findings align with existing literature from other high-income settings, where free STI testing reduces financial barriers but struggle to reach marginalised groups (10). Word-of-mouth and social media effectively attracted young people, students and women, while heterosexual men and adolescents under 18 remained harder to engage (4,11). The observed gender differences reflect prior studies showing women’s greater engagement with preventative care compared to men, who require targeted outreach (12,13).

Consistent with previous implementation research, HCPs reported that the counselling and information-sharing aspects of the intervention were particularly beneficial, not only for individual education but also to diffuse sexual health knowledge through informal social networks (14). A quantitative evaluation of the same pilot supports the importance of the education component, highlighting that people who had a VCT consultation perceived improvements in their sexual health knowledge and valued the non-judgmental discussions about sexuality (7).

Additionally, HCPs noted that early positive experiences with sexual healthcare could reduce stress and encourage individuals to return during future crises. This aligns with literature suggesting that familiarity with healthcare providers and the clinical environment fosters increased health-seeking behaviour and consistent service utilisation (1,15–19), including easier discussions around sensitive topics such as HIV pre-exposure prophylaxis (PrEP) and ongoing engagement with sexual healthcare (20,21).

The perspectives of the HCPs in this study highlight the critical role of free VCT services in promoting equitable access and reducing stigma around sexual healthcare in order to reach national and international goals of STI reduction and HIV elimination (22,23). However, as HCPs noted, access alone is not sufficient. To improve reach, strategies should include mobile testing, culturally tailored outreach, and increased presence in vocational schools and non-academic youth environments.

The findings also suggest the need for broader institutional efforts to normalise sexual health care within mainstream primary care and educational settings. HCPs repeatedly noted the absence of sexual health conversations in general practice and gynaecological care, pointing to a system-wide gap in preventative sexual health. Programmes like this VCT programme can bridge the gap, but sustainable impact will require integration into routine care structures, broader community engagement, and ongoing efforts to destigmatize STI testing across all social groups (3,24,25). HCP offered recommendations focused on increasing visibility, addressing stigma and improving sexual health literacy – suggestions supported by existing evidence (3,12,25,26). Detailed policy recommendations, synthesising these provider insights with current literature, are summarised in Table 2.

**Table 2:** Recommendation for policy makers synthesised from HCP inputs and literature.

| **Focus area** | **Recommendation** | **Literature** |
| --- | --- | --- |
| Financial barriers | Maintain sustained funding for universally free VCT services, especially targeting youth and low-income populations. | (3,4) |
| Stigma and fear discourage testing among conservative and traditional-minded groups | Invest in targeted, community-based interventions among adolescents and emerging adults involving respected community figures or peer-led outreach programmes that address and normalise STI testing in a setting where each person is reached and not associated with stigma such as schools. | (15–17,26,27) |
| Visibility | Increase visibility of sexual healthcare services aimed at young people through strong social media presence aimed at all relevant population groups. | (11) |
| Limited sexual health literacy among young people | Instead of naming the parents as primary source of sexual education, increase the importance of sexual healthcare in schools, but relieve the burden on teaching staff by better publicising all external offers for sex education in schools. | (27,28) |
| Limited sexual health literacy among young people | Introduce opt-out testing for sexually transmitted infections with high burden in target population during gynaecology and general practitioner consultations. | (4,29) |

### Strengths and limitations

Key strengths include diverse HCP representation (counselling, administration, outreach) across both pilot sites, providing comprehensive implementation perspectives. Triangulating qualitative interviews with semi-quantitative activities enriched the analysis. However, limitations include potential reduced generalisability due to single urban location and two testing centres. Using a single interviewer, while ensuring consistency, may have introduced subjective bias during probing. Additionally, social desirability bias may have influenced responses, particularly regarding underreached populations. The findings represent HCPs’ subjective perspectives, an inherent qualitative study limitation.

## Conclusion

Free VCT programmes enhance access to sexual healthcare among young people and first-time testers, introducing many to sexual healthcare services for the first time. However, targeted outreach, structural adaptation, and inclusive communication strategies are essential to expand reach and reduce inequities. Future interventions must specifically target young and low-income groups with culturally and contextually appropriate strategies, integrating deep prevention within schools and communities. These insights are critical for refining and scaling free testing initiatives in Switzerland and other high-income contexts seeking to meet national and global sexual health goals.

## Data Availability

Anonymised excerpts of the interview data that support the findings of this study are available from the corresponding author upon reasonable request. Full transcripts are not publicly available due to privacy and ethical restrictions.

## Acknowledgements

We sincerely appreciate all HCPs who agreed to be interviewed for the scientific evaluation of the pilot project for their time and insights, as well as the clients sharing their data for scientific evaluation. We would like to thank Christina Grube for testing the interview guide. Furthermore, we would like to acknowledge the City of Zurich for funding the scientific evaluation of the free testing pilot project.

## Authors’ Contributions

BH and JSF conceived the initial idea for the scientific evaluation of the pilot project, LAG and AF planned the study, BH organised the recruitment of the HCPs, LAG conducted the interviews, EW and LAG transcribed and coded the interviews. VN refined the themes based on emerging concepts. AF, EW and LAG carried out the data frame analysis. LAG and AF wrote the first draft. All authors read and critically revised the manuscript.

## Declarations

### Patient and public involvement

Patients and/or the public were not involved in the design, conduct, reporting or dissemination plans of this research.

### Patient Consent for Publication

Not applicable

### Ethics Approval and Consent to Participate

The study has been exempted from ethical approval under the Human Research Act by the Cantonal Ethics Committee of Zurich (Req-2023-00594) as data were collected anonymously.

