## Supplemental Material for "Reaching young and low-income people for sexual healthcare – perspectives from healthcare workers: a qualitative study"

**Title:**

Reaching young and low-income people for sexual health care – perspectives from health care workers

**Contents:**

**Supplementary Material S1:** SRQR Checklist

**Supplementary Material S2:** Interview Guide for Health Care Providers’ Interview

**Supplementary Material S3:** Characterisitcs and Potential Influences of Interviewer

**Supplementary Material S4:** Semi-Quantitative Tasks

|  | Item Description | Location (or reason for not reporting) |
| --- | --- | --- |
| **Title & Abstract** |  |  |
| [Title](https:/resources.equator-network.org/reporting-guidelines/srqr/items/title.html) | Describe the nature and topic of the study. Identify the study as qualitative or indicate the approach or data collection methods. | Title page |
| [Abstract](https:/resources.equator-network.org/reporting-guidelines/srqr/items/abstract.html) | Summarise the key elements of the study using the abstract format of the intended publication. | Lines 1-27, page 2 |
| **Introduction** |  |  |
| [Problem Formulation](https:/resources.equator-network.org/reporting-guidelines/srqr/items/problem-formulation.html) | Describe the problem/phenomenon studied, its significance, relevant theory and empirical work, and gaps in current knowledge. | Paragraph 4, lines 59-65 |
| [Purpose or research question](https:/resources.equator-network.org/reporting-guidelines/srqr/items/purpose.html) | Describe the purpose of the study and specific objectives or questions. | Paragraph 4, lines 63-68 |
| **Methods** |  |  |
| [Qualitative approach and research paradigm](https:/resources.equator-network.org/reporting-guidelines/srqr/items/qualitative-approach.html) | Describe your qualitative approach, your guiding theory (if appropriate), and research paradigm, and reasons for your choices. | Paragraph 5, lines 71-76 |
| [Researcher characteristics and reflexivity](https:/resources.equator-network.org/reporting-guidelines/srqr/items/researcher-characteristics-and-reflexivity.html) | Describe how researchers’ characteristics may influence the research, including personal attributes, qualifications/experience, relationship with participants, assumptions, and/or presuppositions; potential or actual interaction between researchers’ characteristics and the research questions, approach, methods, results and/or transferability. | Supplemental Materials S3 |
| [Context](https:/resources.equator-network.org/reporting-guidelines/srqr/items/context.html) | Describe the setting/site(s) in which the study was conducted, why it was selected, and any other salient contextual factors that may influence the study. | Paragraph 6, lines 78-83 |
| [Sampling strategy](https:/resources.equator-network.org/reporting-guidelines/srqr/items/sampling-strategy.html) | Describe how and why research participants, documents, or events were selected; criteria for deciding when no further sampling was necessary, and the rationale for those criteria. | Paragraph 7, lines 87-89 |
| [Ethical issues pertaining to human subjects](https:/resources.equator-network.org/reporting-guidelines/srqr/items/ethics.html) | Describe any approval by an appropriate ethics review board and participant consent, or explain any lack thereof. Describe any other confidentiality and data security issues. | Paragraph 12, lines 126-132 |
| [Data collection methods](https:/resources.equator-network.org/reporting-guidelines/srqr/items/data-collection-methods.html) | Describe the types of data collected; details of data collection procedures including (as appropriate) start and stop dates of data collection and analysis, iterative process, triangulation of sources/methods, and modification of procedures in response to evolving study findings. Describe your rationale for these choices. | Paragraph 8, lines 96-110 |
| [Data collection instruments and technologies](https:/resources.equator-network.org/reporting-guidelines/srqr/items/data-collection-instruments.html) | Describe any instruments (e.g., interview guides, questionnaires) and devices (e.g., audio recorders) used for data collection; describe if/how the instrument(s) changed over the course of the study. | Paragraph 8, lines 95-110  Supplement S2, Supplemental Materials S3 |
| [Units of study](https:/resources.equator-network.org/reporting-guidelines/srqr/items/units-of-study.html) | Describe the number and relevant characteristics of participants, documents, or events included in the study. Describe the level of participation. | Paragraph 7, lines 90-94 |
| [Data processing](https:/resources.equator-network.org/reporting-guidelines/srqr/items/data-processing.html) | Describe the methods for processing data prior to and during analysis, including transcription, data entry, data management and security, verification of data integrity, data coding, and anonymisation / deidentification of excerpts. | Paragraph 8, lines 100-103;  Paragraph 11, lines 116-124  Paragraph 12, lines 128-132 |
| [Data analysis](https:/resources.equator-network.org/reporting-guidelines/srqr/items/data-analysis.html) | Describe the process by which inferences, themes, etc. were identified and developed, including the researchers involved in data analysis; usually references a specific paradigm or approach. Describe why you chose this process. | Paragraph 11, lines 116-124 |
| [Techniques to enhance trustworthiness](https:/resources.equator-network.org/reporting-guidelines/srqr/items/trustworthiness.html) | Describe any techniques to enhance trustworthiness and credibility of data analysis,(e.g., member checking, triangulation, audit trail). Describe why you chose these techniques. | Paragraph 9, lines 104-110  Paragraph 11, lines 116-124 |
| **Results** |  |  |
| [Synthesis and interpretation](https:/resources.equator-network.org/reporting-guidelines/srqr/items/synthesis-and-interpretation.html) | Describe the main findings (e.g., interpretations, inferences, and themes); might include development of a theory or model, or integration with prior research or theory. | Paragraphs 13 – 26, lines 134-215 |
| [Links to empirical data](https:/resources.equator-network.org/reporting-guidelines/srqr/items/links-to-empirical-data.html) | Provide evidence (e.g., quotes, field notes, text excerpts, photographs) to substantiate analytic findings. | Paragraph 13, lines 138-139; paragraph 16, lines 154-156; paragraph 20, lines 172-173; paragraph 23, lines 193-194, table 1, figure 1 |
| **Discussion** |  |  |
| [Integration with prior work, implications, transferability, and contribution(s) to the field](https:/resources.equator-network.org/reporting-guidelines/srqr/items/integration-with-prior-work.html) | Summarize the main findings, explain how findings and conclusions connect to, support, elaborate on, or challenge conclusions of earlier scholarship; discuss the scope of application/generalizability; identify unique contribution(s) to scholarship in a discipline or field. | Paragraphs 27-31, lines 217-273  Table 2 |
| [Limitations](https:/resources.equator-network.org/reporting-guidelines/srqr/items/limitations.html) | Discuss the trustworthiness and limitations of findings | Paragraph 32, lines 268-273 |
| **Other** |  |  |
| [Conflicts of interest](https:/resources.equator-network.org/reporting-guidelines/srqr/items/conflicts-of-interest.html) | Describe any potential sources of influence or perceived influence on study conduct and conclusions. Describe how these were managed. | Paragraph 38, lines 302-304 |
| [Funding](https:/resources.equator-network.org/reporting-guidelines/srqr/items/funding.html) | Describe sources of funding and other support. Describe the role of funders in data collection, interpretation, and reporting. | Paragraph 36, lines 294-297 |

**Supplemental Materials S2: Interview Guide Health Care Providers**

**Pre-Interview (Checklist for Interviewer)**

*The interview takes place in a quiet environment. Open questons will be asked, the interviewee will be encouraged to talk, follow-up questions are asked openly. The flow of conversation will not be interrupted.*

- Welcoming and introduction: I am XX and I work at the EBPI of the University of Zurich and would like to welcome you to the interview, that I will guide you through. Thank you very much for taking the time for this interview. We are very grateful to you, since your view are very important to the project and we know how busy you are. The interview will last no longer than an hour.
  - *I will ask you mostly open-ended questions. You answer whatever spontaneously comes to your mind and what you want to tell me.*
  - *With your permission, I would like to record this interview, so none of your answers get lost, I will be making additional notes and with your permission I would like to photograph the final sorting of the semi-quantitative activities (of the order of cards, you will not be visible). All information (sound, writing, images) will be treated confidentially and will not be marked with your name. Do you agree to be interviewed? Do you agree to the interview to be recorded?*
  - *You can stop the interview at any time or not answer questions. However, I ask you to be as honest and precise in answering the questions you are comfortable to address.*
- *Do you have any questions so far (leave enough time for the interviewee to think about it).*

**Introduction to the interview**

• There are many reasons why sexually active people do not get tested regularly for HIV and STIs. However, valid barriers can be defined for the majority.

• High costs are one of these barriers.

•With this project, we are trying to remove this barrier for two vulnerable population groups, namely those under the age of 26 and people who hold a KulturLegi card.

•We would like to have this conversation to find out about your experiences, whether we have effectively narrowed down the population group, or whether you think that other people are not getting tested for financial reasons despite the existing risk.

•However, we would also like to hear your general thoughts and opinions on the project.

I will now ask you some open-ended questions, and you can simply tell me what you think is important and what you would like to share.

**Do you have any questions at this point? (Wait long enough)**

Ok, so before we start the interview I am asking you to fill in this form:

Prename

Age

Education

Occupation

Job percentage

Further employments

Thank you for filling out the form. If you don’t have any further questions, I would start with the recording and the interview.

| **Barriers** | |
| --- | --- |
| **Sorting Activity of Barriers**  In the intro we have talked about barriers one could face when trying to get tested. Please, rank these barriers in the order of the importance to the populations qualifying for the project as if the project does not exist. | See table 1 for list of barriers |
| Questions during activity:  *Generally, why do you think sexually active people, with a medium to high exposure risks don’t get tested although there are several options outside of GP clinics in Zurich?* | *Get assessment for the different barriers:*  *For whom is barrier relevant?*  *On which observations do you base this assessment?*  *How do they get to know about people who want to come testing but don’t come?* |
| What do you think: Would the expansion of the programme to people up to 30 be doable within the infrastructure? | Do you think such an expansion would make sense?  If no, why not? Would it make more sense to include another population group? If yes, which group(s)?  Can you tell me something about how you came to these assessments? |
| **Tester Demographics** | |
| **Step 1**  We have a scale of need for VCT here on the sheets of paper. This exercise is not about each person in this group having that risk but just a very generalised average. Can you rank these categories in a way that makes the most sense to you. There can be more than one group per step. After the initial sorting, if you want you can add additional defining charateristics to the main group (i.e. “people with little knowledge of safer sex”, addition of “who have multiple partners”). If you feel like a group cannot be ranked or you don’t know or want, you can skip groups.  **Step 2**  How do you define the need for VCT for group X?  **Step 3**  Which of the groups in the two groups of the highest need do you hardly see or never see here?  **Step 4**  Do you have any ideas or suggestions hwo we could reach these people you marked at high need but you don’t see at clinic? | See tbale 2 for list of groups  Age groups  Minorities  Exposure groups  Do you feel that there were changes in people who come or who don’t come anymore? If yes, who comes more and who less? |
| **Nutzen des Pilotprojekts** | |
| Do you think the pilot project is useful? | Do we reach the right people?  Is it efficiently organsied (is there a lot of idle time)?  Are many people coming with low risk who also don’t profit from the counselling? |
| What do you think testers appreciate the most from the project? | Test results  Judgment-free talk about sexuality  Sex ed / Information |
| How could the project be improved? | Procedures  People we should include or exclude… |
| Now we have mainly talked about the positive side of the project for the testers In your opinion, are there negative consequences for the testers? | Is there peer pressure for getting tested?  Are the test takers struggling with sever anxiety about the results?  Do test takers feel stigmatised?  Do you think there is an over treatmnet of infections? |
| **Time management – only counsellors** | |
| Currently, are consultations taking longer or shorter than planned? | What characterises consultations lasting shorter/longer than planned?  How could the planning be improved? |
| How would you evaluate the ratio between counselling and administrative workload (i.e. data entry)? | What would help you better manage the administrative part of your work?  Would there be anything else besides reducing the number of consultations?  How is it working with BerDa in daily routine? |
| **Time management only Intake staff** | |
| How has your work changed since the project started in June?  Do people have to be turned away who don't meet the criteria (over 26, no KL, not from the city)? | Were there incidents where people had to be turned away?  Are there changes regarding waiting times etc.?  Is the number increasing or decreasing?  How is it for you to turn people away?  Have there been problems with this?  Can the tests be financed differently if the tests are really necessary and the person cannot afford them? |
| **End of Interview** |  |
| I have asked all my questions. Do you have something more you want to add or any questions?  Thank you very much for the conversation and for your openness. |  |
| Thank for the talk and «farwell» to interviewee |  |

**Supplemental Materials S3: Characteristics of Interviewer and their Potential Influences on Study**

**Researcher Characteristics:** The primary researcher conducting interviews was a 34-year-old, heterosexual, Western European, cisgender woman with >5 years experience in public health research. However, this was her first qualitative study.

**Relationship with Participants:** The researcher met several health care providers (HCPs) prior to the interviews in meetings on implementing the scientific evaluation on the free testing and had e-mail correspondence with the HCPs overseeing data entry.

**Assumptions and Presuppositions:** Prior to data collection and maintained throughout analysis, the researcher held several assumptions about the programme's effectiveness and reach. These beliefs, documented before coding and analysis, included:

- The programme would be fundamentally beneficial with most clients deriving value from participation.
- Many participants would present with low STI/HIV risk but gain significant benefit from the counselling component.
- The programme would primarily attract heteronormative individuals seeking first-time testing and women who receive limited sexual health guidance from gynecologists.
- Less socially connected individuals would be less likely to learn about the programme.
- Individuals with higher education and flexible work schedules would be overrepresented due to easier appointment access.

**Methodological Safeguards:** To minimise potential researcher influence, the study employed several strategies. The interview guide was developed collaboratively with other researchers and piloted prior to data collection to ensure consistency and reduce individual bias. The coding system was developed jointly with two other researchers, and framework analysis was conducted with two team members to enhance analytical rigor and reduce individual interpretation bias.

**Potential Influence on Research:** Despite these safeguards, the researcher's assumptions and demographic characteristics may have influenced interview questioning, data interpretation, and the emphasis placed on themes related to programme accessibility and reach among different population groups.

**Supplemental Material S4: Semi-quaqntitative tasks**

Barriers

Opening hours

Costs

Shame / Embarassement

Lack of education / information

Fear of results

Fear of tests

Lack of knowledge on testing sites

Fear of reaction from social environment

No motivation

Time constraints

Long waiting periods for appointment

Table 2

| Population group categories | |
| --- | --- |
| Students | Asylum seekers |
| No formal education | Short stay in Switzerland |
| Vocatinoal students | Long-term resident Switzerland / Swiss citizen |
| < 18 year olds | Sex workers |
| 18 – 25 year olds | Eastern Europe |
| 26 – 35 year olds | Africa |
| 36 – 45 year olds | Asia |
| > 45 year olds | Oceania |
| Tourists | North America |
| Urban | Latin America |
| Metropolitan area | Moderate knowledge on safer sex |
| Rural | Good knowledge on safer sex |
| Little knowledge on safer sex |  |
